# Motor imagery interventions in stroke patients modulate their brain-heart network dynamics

**DOI:** 10.64898/2026.06.23.26355958

**Authors:** Diego Candia-Rivera, Marie-Constance Corsi, Juliana Gonzalez-Astudillo, Charlotte Rosso, Fabrizio de Vico Fallani, Mario Chavez

## Abstract

**Background:** Motor imagery is widely used in post-stroke rehabilitation research, as it is thought to promote neuroplastic mechanisms underlying motor recovery. The characterization of the associated neurophysiological patterns is therefore crucial to improve our understanding and eventually optimize rehabilitative brain–computer interface applications. Emerging evidence suggests that cardiac dynamics are actively related to motor-related brain processes, yet the role of brain–heart interactions (BHI) in post-stroke recovery remains largely unexplored. This study aimed to investigate how BHI evolve during motor imagery interventions in stroke patients, with the hypothesis that BHI would exhibit a progressive change across motor imagery interventions.

**Methods:** We analyzed two independent cohorts comprising a total of 15 stroke survivors with upper or lower limb motor impairments undergoing motor imagery-based interventions. BHI were quantified by assessing the coupling between cardiac sympathetic activity and EEG-derived brain network metrics, including clustering and assortativity, across multiple sessions. We examined BHI in three different contexts: i) when motor imagery was combined with electrical stimulation, ii) throughout longitudinal motor imagery interventions, and iii) across longitudinal assessments of behavioral changes.

**Results:** Motor imagery combined with electrical stimulation on the affected limb modulated BHI across a wide range of frequencies. In longitudinal interventions, BHI showed a progressive increase, specifically reflected in stronger coupling between the cardiac-sympathetic index (CSI) and beta-band clustering. Moreover, changes in functional independence were tracked by variations in the coupling between CSI and EEG gamma-band networks assortativity.

**Conclusions:** Our findings suggest that BHI dynamically reorganize during post-stroke recovery and are modulated by motor imagery interventions. The observed change in BHI suggests that these interactions may reflect underlying neuroplastic processes. As such, BHI represent a promising class of multisystem biomarkers for tracking recovery and could inform the development of more adaptive and physiologically grounded BCI-based rehabilitation strategies.

## Background

Motor imagery has emerged as a promising tool for motor rehabilitation and brain–computer interfaces (BCIs) in individuals with post-stroke motor impairments [1–4]. By engaging neural circuits involved in action without requiring overt movement, motor imagery offers a valuable alternative when motor execution is limited or absent [4]. However, stroke does not only affect focal brain regions but also disrupts large-scale physiological integration [5], altering systemic mechanisms that support motor imagery [6]. This disruption may contribute to the substantial inter-individual variability observed in motor imagery performance and to the inconsistent efficacy of motor imagery-based rehabilitation strategies [7,8]. Understanding the physiological substrates of motor imagery in stroke patients is therefore critical for improving both clinical interventions and BCI applications [9].

Detecting the signatures of covert motor intent has enabled advances ranging from assistive BCIs to the assessment of residual awareness and motor function [10,11]. However, in stroke populations, these mechanisms may be profoundly altered due to lesion-induced reorganization and compensatory plasticity. Although previous work has shown that motor imagery engages distinct components of the motor cortex [12,13], its underlying mechanisms from a systemic perspective remain incompletely understood, especially in pathological conditions. Confounding factors such as lesion location, severity, and alterations in network connectivity likely contribute to variability in motor imagery ability, while robust non-invasive methods for capturing these altered mechanisms remain challenging.

Various studies have shown that motor imagery can engage both brain and peripheral physiological activity [14–18], including the brain–heart axis [19,20]. These brain–heart interactions are context-dependent and may reflect changes in task demands, learning, and functional reorganization.

Stroke is recognized as a disorder that extends beyond focal brain damage and affects the communication between the brain and others organs [21]. Clinical and experimental studies have demonstrated that stroke can disrupt autonomic regulation, alter heart rate variability, impair cardiovascular control, and increase the risk of cardiac complications [22–28], reflecting a profound disturbance of brain–heart interactions. These alterations have been associated with poorer functional outcomes, reduced recovery potential, and increased mortality [29]. Despite growing evidence that brain–heart dysregulation is a key feature of stroke pathology, the underlying electrophysiological interactions between these organs remain unexplored. Few studies have directly characterized how stroke affects the dynamic exchange of information between neural and cardiac signals, limiting our understanding of the mechanisms through which brain–heart communication contributes to recovery, functional impairment, and rehabilitation outcomes. Recently, research has shown that cardiac reflexes can index impaired blood flow velocity in major cerebral arteries [30], and also that post-stroke cognitive impairment is associated with disrupted brain–heart coupling, particularly during cognitive load, and that these disruptions are related to cognitive impairment severity [31]. Investigating these interactions is especially relevant given emerging evidence that interventions targeting the autonomic nervous system, such as vagus nerve stimulation, can enhance motor recovery after stroke [32]. The reported benefits of vagus nerve stimulation suggest that modulation of brain–heart pathways may influence neuroplasticity and rehabilitation outcomes, further highlighting the need to elucidate the electrophysiological mechanisms linking cardiac and neural function in stroke patients.

We hypothesized that brain–heart interactions are modulated during motor imagery in stroke patients. We expected that stroke-related alterations would manifest as changes in the organization of brain–heart dynamics, potentially indexing compensatory mechanisms or maladaptive processes. To test these hypotheses, we studied longitudinal motor imagery interventions in stroke patients, in which participants performed a motor imagery task of upper or lower limb movements, depending on the specific affection of the patient. This paradigm allowed us to quantify the physiological modulations across motor imagery sessions. We applied a recently developed framework to assess brain–heart interplay by relating brain network organization to indices of cardiac sympathetic activity [33].

## Materials and methods

### Participants

#### Dataset 1

Dataset 1 corresponds to an open access resource, gathered from Figshare [34]. This dataset comprises stroke patients, confirmed by head CT or MRI, with hemiplegia. Patients had a disease duration of 1–12 months and age of 30–70 years. Exclusion criteria included motor dysfunction caused by other diseases, significant cognitive, language, or communication impairments, inability to place their heels on the ground, and severe cardiac conditions, including implanted cardiac pacemakers.

From a total of 27 patients recruited, we studied a sub-cohort of 9 (see Table 1) based on their ECG availability (see Data pre-processing section).

**Table 1.**
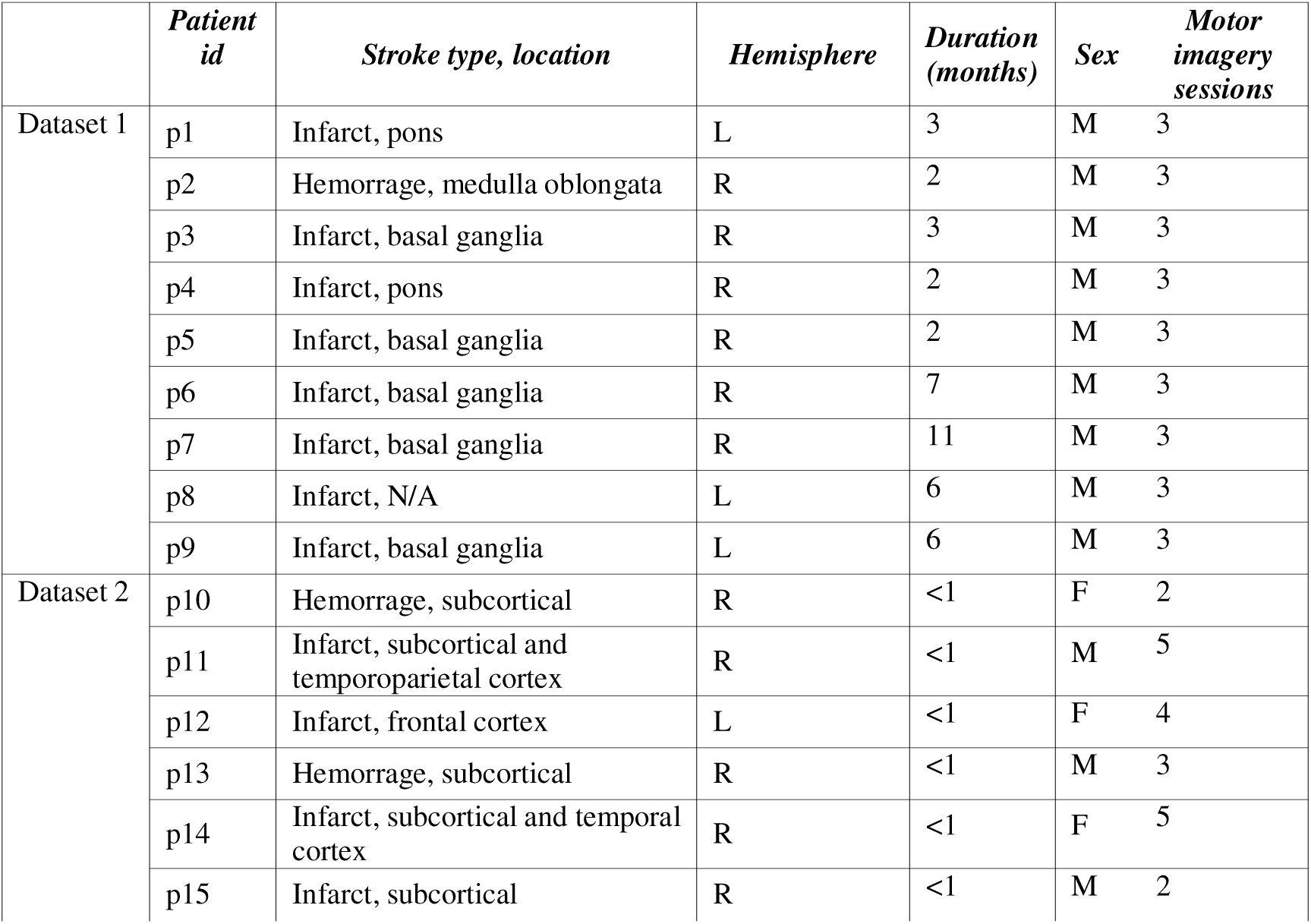
Demographic and clinical details of the cohort.

**Table 2.** Non-parametric, paired sign rank tests comparing brain-heart interactions under motor imagery against motor imagery coupled with electrical stimulation (from Dataset 1), either invariant electrical stimulation (IES) or synchronous electrical stimulation (SES).

|  | Motor Imagery vs (Motor imagery + IES) |  |  | Motor Imagery vs (Motor imagery + SES) |  |  |
| --- | --- | --- | --- | --- | --- | --- |
|  | Alpha | Beta | Gamma | Alpha | Beta | Gamma |
| CSI–Clustering | Z = -1.71781<br>p = 0.103 | Z = -1.48087<br>p = 0.13864 | **Z = -2.66557**<br>**p = 0.02172** | **Z = -2.66557**<br>**p = 0.00769** | **Z = -2.66557**<br>**p = 0.00769** | **Z = -2.66557**<br>**p = 0.00769** |
| CSI–Assortativity | **Z = -2.54710**<br>**p = 0.02172** | **Z = -2.42863**<br>**p = 0.02274** | **Z = -2.66557**<br>**p = 0.02172** | **Z = -2.66557**<br>**p = 0.00769** | **Z = -2.66557**<br>**p = 0.00769** | **Z = -2.66557**<br>**p = 0.00769** |

*Bold indicates significant at FDR corrected $\alpha = 0.05$*

#### Dataset 2

Dataset 2 corresponds to locally acquired data. This dataset comprises stroke patients, confirmed by head MRI, with hand motor impairment. Patients had a disease duration of 5–14 days and age of 44–80 years. Exclusion criteria included significant cognitive, language, or communication impairments.

A total of 6 post-stroke patients were recruited (see Table 1).

### Protocol

#### Dataset 1

The task consisted in a visually guided gait motor imagery, in which patients were informed about the gait phase they had to follow. Patients watched an example of gait task video to familiarize themselves with the paradigm prior data acquisition. Each trial consisted of approximately 5 minutes, including forty 5-second runs, randomly sorted as motor imagery or rest. The experiment was performed in a longitudinal design of three sessions. Session 1 included two additional motor imagery experiments, with invariable electrical stimulation (IES) and a sequential electrical stimulation (SES) following the gait phase, both in the affected leg. Session 2 occurred 2–4 weeks after session 1. Session 3 occurred 4–6 weeks after session 2.

Data collection was performed using a NeuSenW amplifier (Neuracle, Inc.) with a 64-channel EEG sampled at 1000 Hz. For this dataset, cardiac dynamics were reconstructed indirectly from EEG recordings through ICA-based extraction of ECG-related components, as per previous studies [35].

#### Dataset 2

The protocol consisted in motor imagery of the affected hand, guided by a screen where a hand was displayed to instruct the moments of motor imagery. The motor imagery condition consisted in hand grasping, where patients imagined closing their fist to grab an object and raise it upwards. Patients were seated with palms facing upward in front of a screen. Each trial consisted of approximately 2 minutes, including 12 runs, randomly sorted as motor imagery or rest. The experiment was performed with a longitudinal design of five visits over the year following the stroke. The first session was within the first 10 days after the stroke. The following session were performed at 1, 3, 6 and 12 months after the stroke.

Patients were evaluated using a standardized clinical assessment protocol. Functional and neurological status were measured with the Modified Rankin Scale (mRS) and the National Institutes of Health Stroke Scale (NIHSS). Independence in daily living activities was assessed using the Functional Independence Measure (FIM) and the Barthel Index. Motor recovery was quantified with the Fugl-Meyer Assessment. All evaluations were performed by trained clinicians following established scoring guidelines.

Data collection was performed using a BrainAmp EEG system (Brain Products GmbH) with a 64-channel coupled with ECG acquisition, pre-amplified and sampled at 500 Hz.

### Data preprocessing

Physiological signals were processed using MATLAB R2024b and Fieldtrip Toolbox [36]. The EEG and ECG data were bandpass filtered within 0.5–45 Hz using a Butterworth filter of order 4. Large movement artifacts were removed from EEG using a wavelet thresholding on the independent component analysis (ICA) [37]. After large artifacts removal, ICA was run to detect physiological artifacts, including eye movements and the cardiac field artifact. Components identified as containing physiological artifacts were excluded from the EEG reconstruction. In Dataset 1, the ECG was not recorded; therefore, during the ICA step, the component containing heartbeat activity was retained and used as a proxy ECG signal. This approach was successful in 9 out of 27 patients (see Table 1). In contrast, Dataset 2 included ECG recordings, which were incorporated into the ICA computation to improve the identification and removal of cardiac artifacts. After denoising, EEG channels were finally re-referenced using a common average [38].

The R-peaks from the ECG were identified using an automatized process based on the Pan-Tomkins algorithm, followed by an automated inspection of misdetections and a visual confirmation. Recordings that required manual corrections were further processed using R-DECO toolbox [39]. Manual corrections were performed with a visual inspection over the original ECG and the R-R tachogram.

Brain-heart interaction analysis follows a previously developed pipeline [33], that follows the computation of time-resolved brain network dynamics, cardiac indices and their coupling on time (Figure 1).

**Figure 1.**
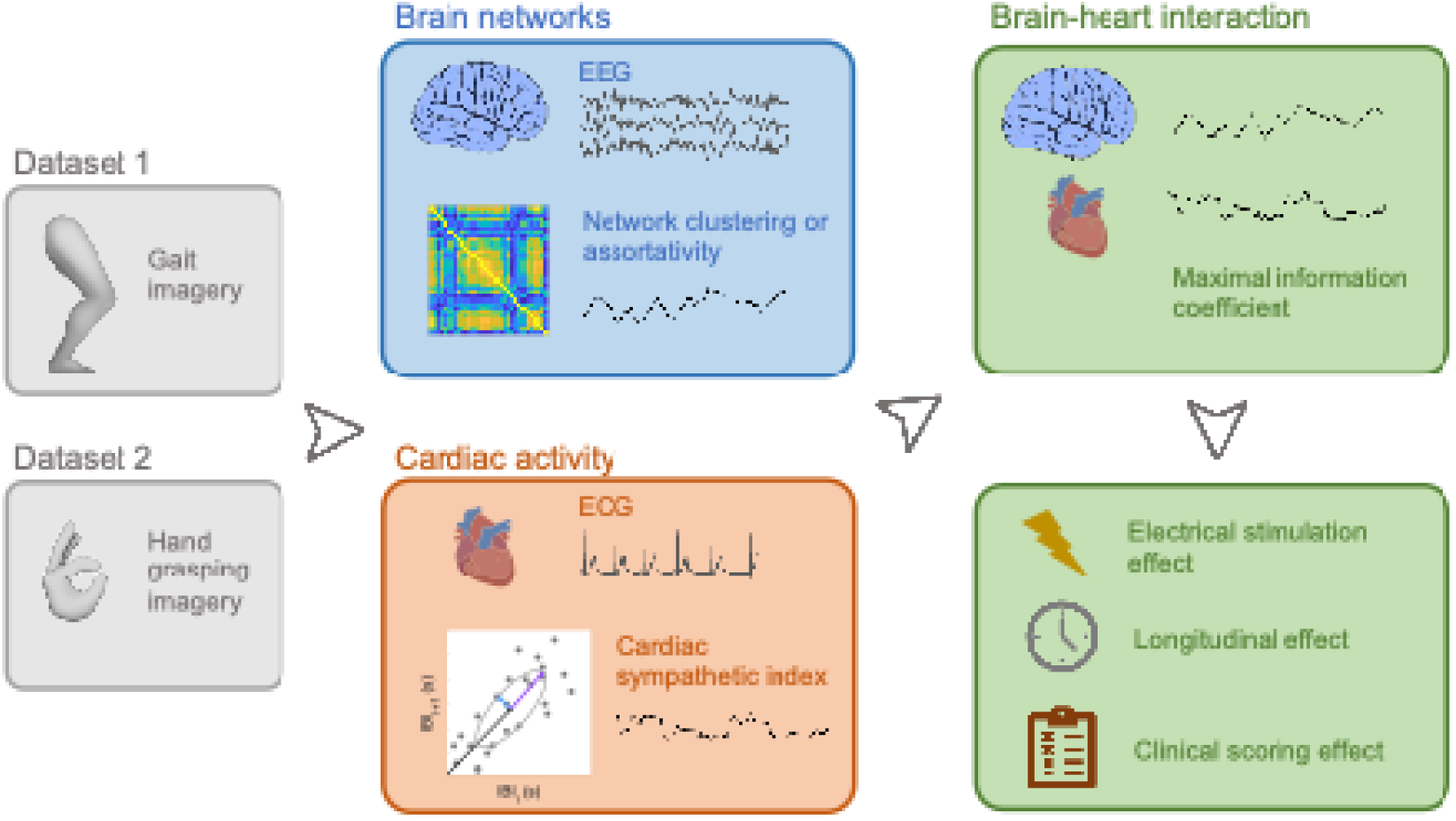
Experimental procedure and data analysis pipeline. EEG and ECG recordings from Datasets 1 and 2 were processed to extract time-varying measures of brain network clustering and assortativity from coherence-based functional connectivity in the alpha, beta, and gamma frequency bands, together with a cardiac sympathetic index. Brain–heart interactions were quantified using the Maximal Information Coefficient, which captures both linear and non-linear associations between time series. The resulting brain–heart interaction metrics were subsequently analyzed to assess their relationships with motor imagery performance: (i) under electrical stimulation, (ii) across longitudinal changes over time, and (iii) with clinical assessment scores.

### Brain network dynamics

EEG power and cross spectral densities were computed using the short-time Fourier transform with a multi-taper, with a sliding time window of 2 seconds with a 50% overlap. For each pair of EEG time series χ*_i_*(t) and χ_j_(t), and their respective complex Fourier Transform χ*_i_*[*f*] and χ*_j_*[*f*], the coherence *COH_i,j_* [*f*]at the frequency f is defined as:

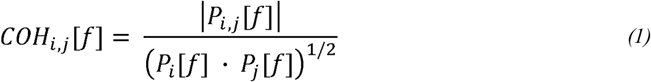

Where P*_i,j_*[*f*] is the cross-spectrum of χ*_i_*[*f*] and χ*_i_*[*f*] and P_n={i,j}_[f] is the power spectral density of χ_n={i,j}_ (t). The output’s absolute value was considered for the network analyses. Coherence matrices were integrated within alpha (8.5–12), beta (12.5–30□Hz) and gamma (30.5–45□Hz) bands, based on previous reports [12,19,40,41]. Connectivity matrices were binarized using a network filter consisting in an efficiency–cost optimization algorithm [42].

Individual connectivity matrices were characterized using global network metrics. This resulted in brain network time series sampled at 1 Hz. We focused in two types of network metrics: clustering and assortativity. These graph-theoretical metrics were retained because they provide complementary information on brain network topology, with clustering coefficient reflecting local functional integration within subnetworks and assortativity describing the preferential connectivity between nodes of similar degree, an important marker of network organization and robustness. Both network metrics were computed using the Brain Connectivity Toolbox [43].

Clustering is a measure that indicates the tendency of the nodes of the network to be grouped, meaning that a high clustering indicates that the networks contains a high amount of groups of nodes, based in their connectivity [44,45]. Clustering was computed as follows:

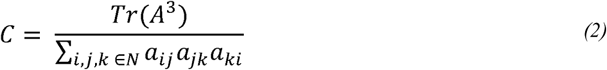

Where A denotes the adjacency matrix, *Tr*is the trace function and *a_i,j_*is connectivity value between the nodes *i*and *j*.

Assortativity is a measure that indicates preference of the nodes of the network to be connected with other nodes with similar connectivity [46]. Assortativity was computed as follows:

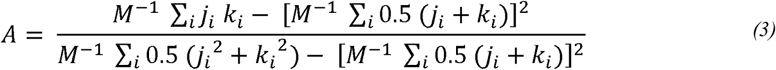

Where *j_i_*, *k_i_* are the degrees of the vertices at the ends of the *i_th_*edge, with = 1, …, *M*.

### Cardiac sympathetic activity estimation

We used our recently developed and validated approach to quantify cardiac rhythms in a time-resolved manner [47–50]. Inter-beat interval (IBI) series were constructed based on the R-to-R-peak durations. Poincaré plot was used to depict the fluctuations on the duration of consecutive IBI [51]. We quantified the heart rate variability component of the cardiac sympathetic index (CSI), which correspond to the long ratios of the ellipsoid representing the long-term fluctuations of heart rate variability [52].

The time-varying fluctuations of the long ellipse ratio was computed with a sliding-time window, as follows:

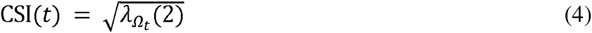

Where *λ_Ωt_* is the vector with the eigenvalues of the covariance matrix of *IBI* _i,…,n-1_and *IBI*_i+1,…,n,_ with Ω_t_: t – T ≤ t_i_ ≤ t and is the length of IBI in the time window Ω_t_. The eigenvalues are ordered in ascending order. Therefore, the second eigenvalue corresponds to the slower rhythm in the defined time window.

Note that the heart rate variability component of CSI is typically correlated with the low frequency (LF) band of the HRV spectrum [47]. It is important to mention that this implementation does not require a frequency band definition, but rather capture the slow and fast rhythms associated with the two computed eigenvalues, that typically fall within LF (<0.15 Hz) and HF (>0.15 Hz).

### Brain-heart network dynamics

For each session, brain-heart interaction analysis was performed over all trials and results were summarized as the median per session and participant.

To quantify the coupling between the fluctuations of brain network metrics and cardiac dynamics we used the Maximal Information Coefficient (MIC). MIC is a method that quantifies the coupling between two time series [53]. MIC evaluates similarities between different segments separately at an adapted time scale that maximizes the mutual information, with a final measure that wraps the similarities across the whole time-course.

The MIC between two time series X and Y was computed as:

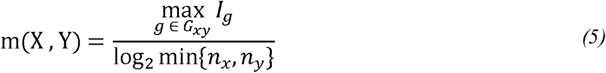

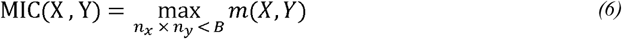

where *G_xy_* denotes the set of all admissible grid partitions of the joint distribution of X and Y, and *I_g_*is the mutual information computed with a particular grid *g*. The quantities represent the numbers of bins used to partition X and Y, respectively, while B is the maximum allowable grid resolution. For each admissible grid, the mutual information values are normalized by the minimum joint entropy log_2_min{n_x_, n_y_}, resulting in a score bounded between 0 and 1. Finally, the quantified coupling between X and Y corresponds to the normalized mutual information resulting from the grid that maximizes the MIC value. The MIC is then defined as the maximum normalized mutual information over all grid partitions satisfying the constraint n_x_. n_y_ < *B* Here, B = N^0.6^, and N is the dimension of the signals [53]. The source code implementing MIC is available online at https://github.com/minepy.

### Statistical analysis

All statistical analyses were conducted to assess how motor imagery combined with electrical stimulation modulates brain–heart interactions across frequencies, how brain–heart interactions evolve throughout longitudinal interventions, and how changes in functional independence and motor impairment relate to brain–heart interaction measures, and their respective counterparts in the brain and heart in isolation.

Modulations of brain–heart interactions during motor imagery coupled with electrical stimulation were assessed using a paired sign rank test, comparing motor imagery vs motor imagery + invariable electrical stimulation, and motor imagery vs motor imagery + sequential electrical stimulation.

Longitudinal effects were examined with linear mixed-effects models. The model aimed to identify brain–heart dynamics consistently modulated by motor imagery. The model was defined as:

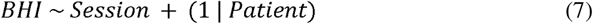

where session was included as an ordered continuous predictor, and patient number as a random effect. Reported p-values correspond to the session effect.

Associations between clinical scores and brain–heart interactions were assessed with linear mixed-effects models. The model aimed to identify whether changes in brain–heart dynamics were associated with changes in the clinical scores. The model was defined as:

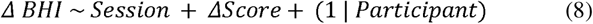

Reported p-values correspond to the score effect. p-values were adjusted using false discovery rate (FDR).

To assess whether patients exhibited a longitudinal improvement in clinical outcome across sessions, we applied Page’s L trend test, a nonparametric test specifically designed for ordered repeated-measures data with an a priori directional hypothesis. For each patient, clinical scores were ranked across sessions while ignoring missing values. An increasing or decreasing trend in clinical scores across sessions (reflecting clinical improvement) was specified as the ordered alternative hypothesis. The Page L statistic was then computed by weighting session-wise ranks according to the hypothesized order. Because of missing observations and the limited sample size, statistical significance was assessed using a permutation-based approach. Within each patient, observed ranks were randomly permuted across available sessions (10,000 permutations), preserving the pattern of missing data. The empirical p-value was calculated as the proportion of permuted L statistics greater than or equal to the observed L statistic.

## Results

### Electrical stimulation effects on brain-heart interactions during motor imagery

We compared brain–heart interaction during motor imagery alone with motor imagery combined with two types of electrical stimulation applied to the affected limb: one with invariant timing and another synchronized with the gait imagery phase. This was performed in the patients from Dataset 1 in their first session.

Electrical stimulation enhanced the coupling between cardiac sympathetic activity and brain network dynamics, reflected in measures of clustering and assortativity. The invariant stimulation mainly affected the CSI–assortativity coupling, whereas the synchronized stimulation influenced both clustering and assortativity couplings (Table□2). Although the modulation of brain–heart interaction was observed across a broad frequency range, the effects were most prominent in the gamma band (Figure□2). Control analyses confirmed that these effects were not significant when testing brain network or cardiac activity changes independently (Table 3).

**Table 3.**
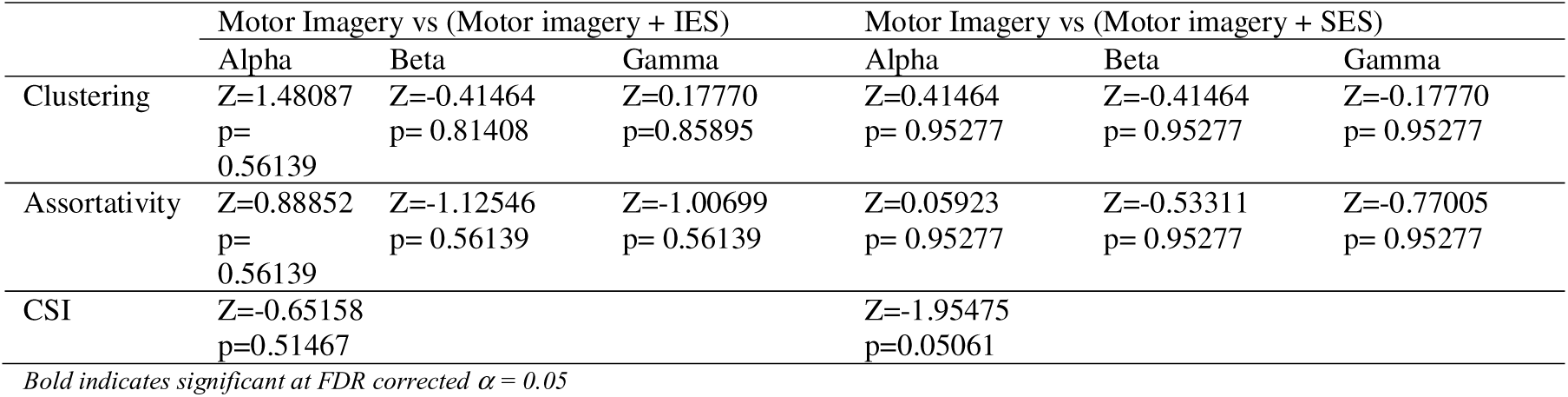
Non-parametric, paired sign rank tests comparing brain network or cardiac changes under motor imagery against motor imagery coupled with electrical stimulation (from Dataset 1), either invariant electrical stimulation (IES) or synchronous electrical stimulation (SES).

|  | Motor Imagery vs (Motor imagery + IES) |  |  | Motor Imagery vs (Motor imagery + SES) |  |  |
| --- | --- | --- | --- | --- | --- | --- |
|  | Alpha | Beta | Gamma | Alpha | Beta | Gamma |
| Clustering | Z=1.48087<br>p=0.56139 | Z=-0.41464<br>p=0.81408 | Z=0.17770<br>p=0.85895 | Z=0.41464<br>p=0.95277 | Z=-0.41464<br>p=0.95277 | Z=-0.17770<br>p=0.95277 |
| Assortativity | Z=0.88852<br>p=0.56139 | Z=-1.12546<br>p=0.56139 | Z=-1.00699<br>p=0.56139 | Z=0.05923<br>p=0.95277 | Z=-0.53311<br>p=0.95277 | Z=-0.77005<br>p=0.95277 |
| CSI | Z=-0.65158<br>p=0.51467 |  |  | Z=-1.95475<br>p=0.05061 |  |  |

Bold indicates significant at FDR corrected $\alpha = 0.05$

### Longitudinal motor imagery modulation of brain–heart interactions

Our analyses revealed significant session-dependent modulations in brain–heart interaction. As shown in Table 4, cardiac sympathetic activity showed a strong and progressive increase in its coupling with beta-band clustering in both datasets (see Fig. 3). A similar effect was found, only in the dataset 1, in the cardiac couplings with clustering and assortativity, both in the alpha-band. A less sustained effect was found in the cardiac couplings with gamma-band clustering and assortativity. These results indicate a strengthening relationship between cardiac sympathetic dynamics and fluctuations in brain network segregation and hub organization over the course of the motor imagery intervention.

**Figure 2.**
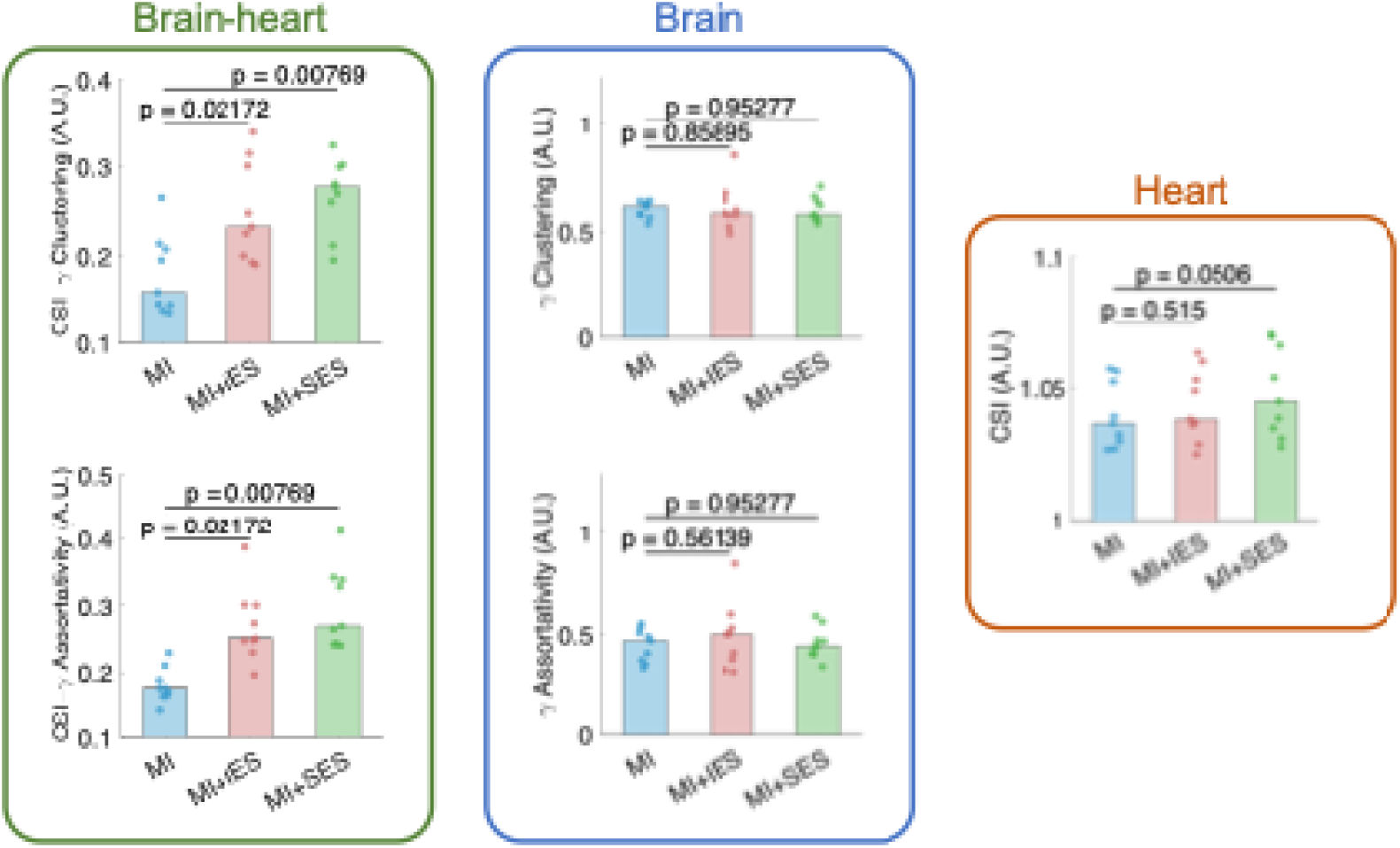
Changes in brain–heart network dynamics during motor imagery combined with electrical stimulation (from Dataset 1). Significant modulations in brain–heart interactions are observed between the cardiac sympathetic index (CSI) and gamma-band clustering and assortativity during motor imagery (MI) when either invariant electrical stimulation (IES) or synchronous electrical stimulation (SES) is applied. A.U., arbitrary units.

**Figure 3.**
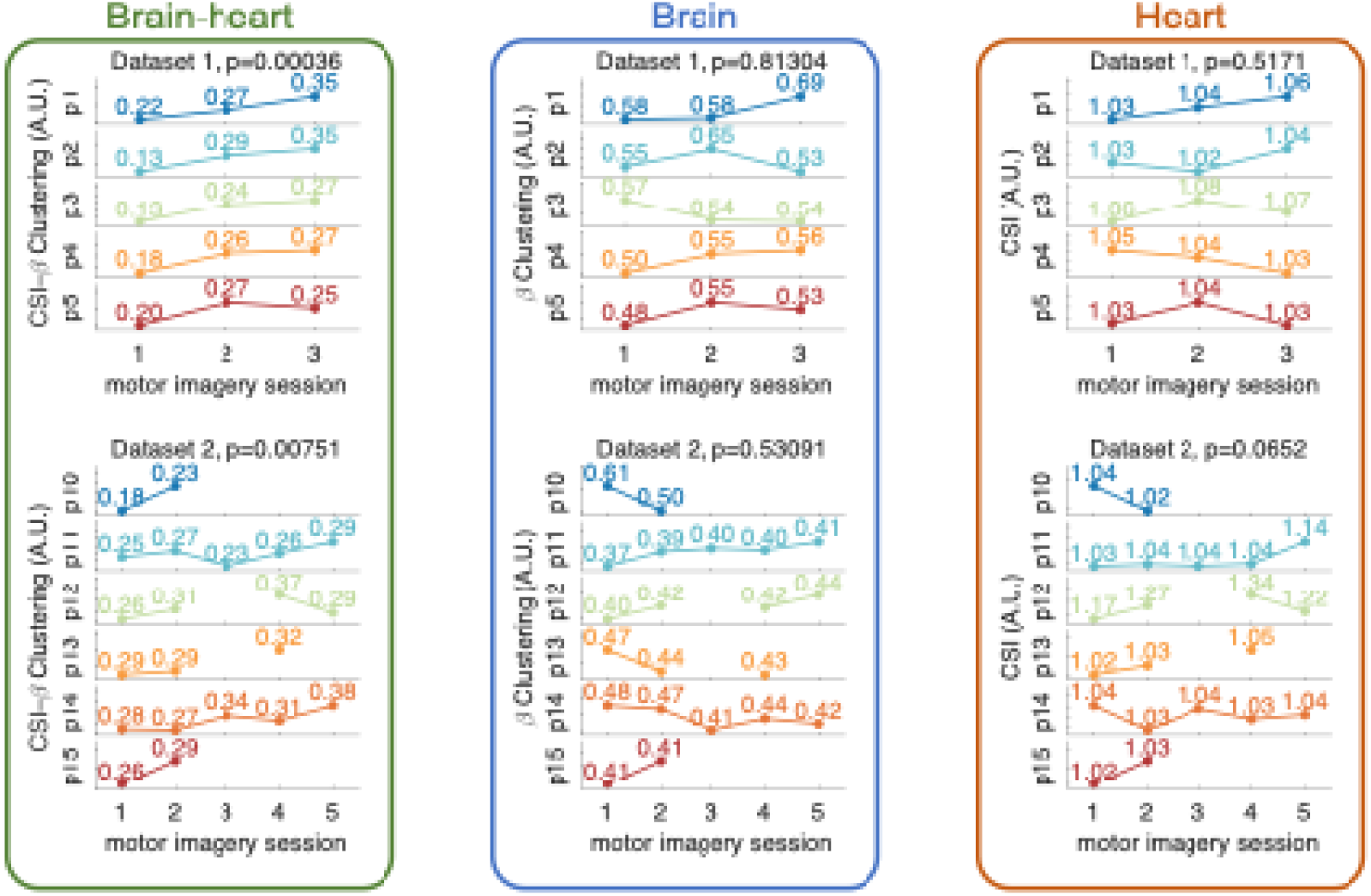
Brain-heart interactions during motor imagery interventions, as quantified from the coupling between beta-band clustering and cardiac sympathetic activity in Datasets 1 and 2. Missing data points mean that the session was not available. A.U.: arbitrary units.

**Table 4.**
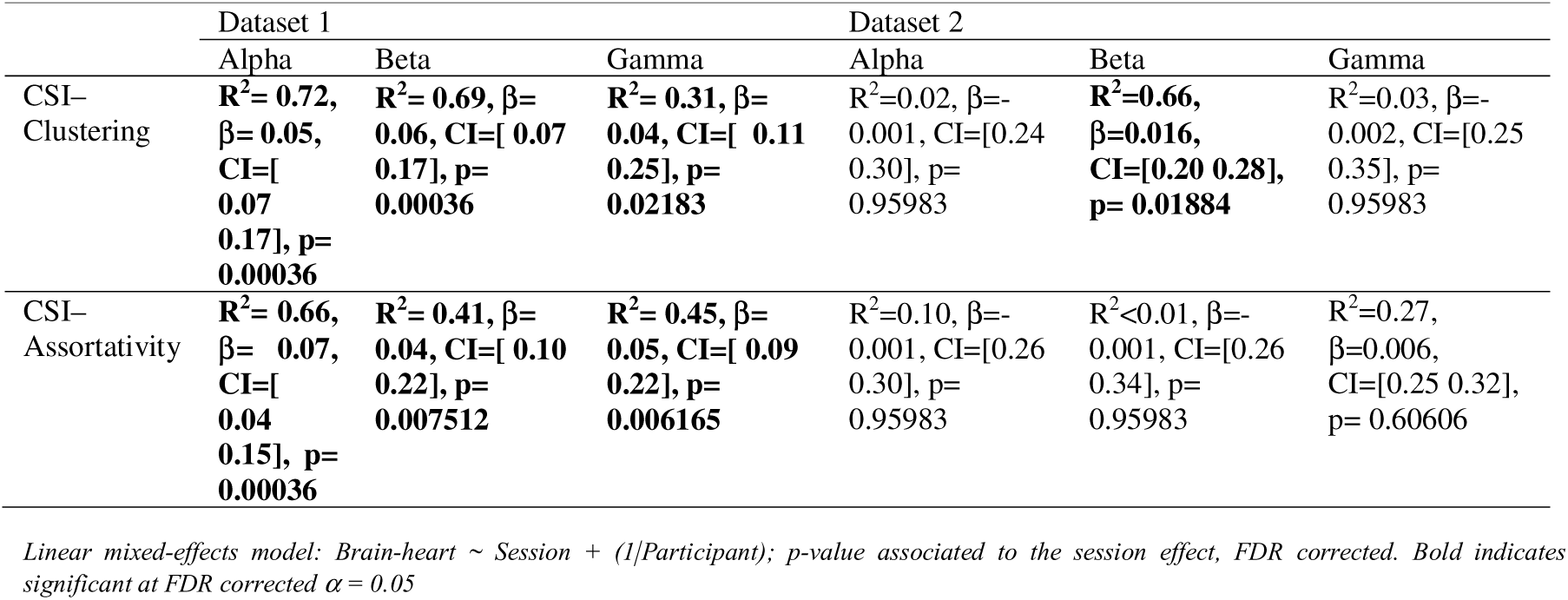
Linear mixed-effect model of the effect of longitudinal motor imagery interventions on brain-heart interactions, in two independent datasets of stroke patients.

|  | Dataset 1 |  |  | Dataset 2 |  |  |
| --- | --- | --- | --- | --- | --- | --- |
|  | Alpha | Beta | Gamma | Alpha | Beta | Gamma |
| CSI-Clustering | $R^2 = 0.72$,<br>$\beta = 0.05$,<br>$CI = [0.07, 0.17]$, $p = 0.00036$ | $R^2 = 0.69$, $\beta = 0.06$, $CI = [0.07, 0.17]$, $p = 0.00036$ | $R^2 = 0.31$, $\beta = 0.04$, $CI = [0.11, 0.25]$, $p = 0.02183$ | $R^2 = 0.02$, $\beta = -0.001$, $CI = [0.24, 0.30]$, $p = 0.95983$ | $R^2 = 0.66$,<br>$\beta = 0.016$,<br>$CI = [0.20, 0.28]$, $p = 0.01884$ | $R^2 = 0.03$, $\beta = -0.002$, $CI = [0.25, 0.35]$, $p = 0.95983$ |
| CSI-Assortativity | $R^2 = 0.66$,<br>$\beta = 0.07$,<br>$CI = [0.04, 0.15]$, $p = 0.00036$ | $R^2 = 0.41$, $\beta = 0.04$, $CI = [0.10, 0.22]$, $p = 0.007512$ | $R^2 = 0.45$, $\beta = 0.05$, $CI = [0.09, 0.22]$, $p = 0.006165$ | $R^2 = 0.10$, $\beta = -0.001$, $CI = [0.26, 0.30]$, $p = 0.95983$ | $R^2 < 0.01$, $\beta = -0.001$, $CI = [0.26, 0.34]$, $p = 0.95983$ | $R^2 = 0.27$,<br>$\beta = 0.006$,<br>$CI = [0.25, 0.32]$, $p = 0.60606$ |

Linear mixed-effects model: Brain-heart ∼ Session + (1|Participant); p-value associated to the session effect, FDR corrected. Bold indicates significant at FDR corrected $\alpha = 0.05$

In contrast, this effect did not generalize across other frequency bands, suggesting a degree of frequency- and topology-specificity in the observed brain-heart network interaction patterns.

We verified whether these associations could be reproduced when analyzing brain and heart metrics separately, confirming that the effect is specific to brain–heart interactions (Table 5).

**Table 5.** Linear mixed-effect model of the effect of longitudinal motor imagery interventions on brain or heart metrics, in two independent datasets of stroke patients.

|  | Dataset 1 |  |  | Dataset 2 |  |  |
| --- | --- | --- | --- | --- | --- | --- |
|  | Alpha | Beta | Gamma | Alpha | Beta | Gamma |
| Clustering | $R^2=0.02$,<br>$\beta=0.003$,<br>CI=[ 0.48<br>0.60],<br>p=0.81304 | $R^2=0.20$,<br>$\beta=0.016$, CI=[<br>0.46 0.60],<br>p=0.81304 | $R^2=0.25$,<br>$\beta=0.005$, CI=[<br>0.53 0.65],<br>p=0.81304 | $R^2=0.41$, $\beta=-$<br>0.009, CI=[0.44<br>0.55],<br>p=0.38566 | $R^2=0.71$, $\beta=-$<br>0.004, CI=[0.41<br>0.52],<br>p=0.53091 | $R^2=0.48$, $\beta=-$<br>0.004, CI=[0.46<br>0.55],<br>p=0.53091 |
| Assortativity | $R^2=0.01$,<br>$\beta=0.003$,<br>CI=[0.32<br>0.47],<br>p=0.81304 | $R^2=0.02$,<br>$\beta=0.013$,<br>CI=[0.30 0.54],<br>p=0.81304 | $R^2=0.11$,<br>$\beta=0.015$,<br>CI=[0.33 0.53],<br>p=0.81304 | $R^2=0.02$, $\beta=-$<br>0.004, CI=[0.37<br>0.45],<br>p=0.53091 | $R^2=0.57$, $\beta=-$<br>0.019, CI=[0.37<br>0.48],<br>p=0.05346 | $R^2=0.37$, $\beta=-$<br>0.026, CI=[0.39<br>0.53],<br>p=0.05346 |
| CSI | $R^2=0.43$, $\beta=0.003$, CI=[1.02 1.06], p=0.51707 | | | $R^2=0.86$, $\beta=0.012$, CI=[0.96 1.13], p=0.06516 | | |

Linear mixed-effects model: Brain or heart ~ Session + (1|Participant); p-value associated to the session effect, FDR corrected. Bold indicates significant at FDR corrected $\alpha = 0.05$

### Brain–heart interactions and functional outcomes

Patients from dataset 2 were admitted with slight disability to moderately severe disability, confirmed with the modified Rankin scale, ranging between 2–4, and with normal to moderate neurological impairment, confirmed with the NIH stroke scale <15. Both scales remained moderately stable within the 1 year (Page tests; mRS: L=178, p=0.0308; NIHSS: L 181, p = 0.0117). We tested whether these brain–heart interactions could also explain functional outcome evolution within 1 year. These tests focused on measures of functional independence in daily life activities (Barthel and MIF scores) and motor impairment (Fugl-Meyer score). First, we observed substantial improvements in both daily life activities tests (Page test; Barthel: L=170, p<0.0001; FIM: L=168, p=0.0002) and motor impairment (Page test, Fugl-Meyer: L=169, p<0.0001). Then, we tested separately per each of these scores which brain–heart network metric better capture those changes. We found that changes in FIM scores can be tracked by the changes in the coupling between CSI and gamma assortativity (Table 6 and Figure 4). We did not find any significant relationship in Barthel of Fugl-Meyer scores. We then confirmed that brain and heart metrics separately do not track any of the clinical scores studied (Table 7).

**Figure 4.**
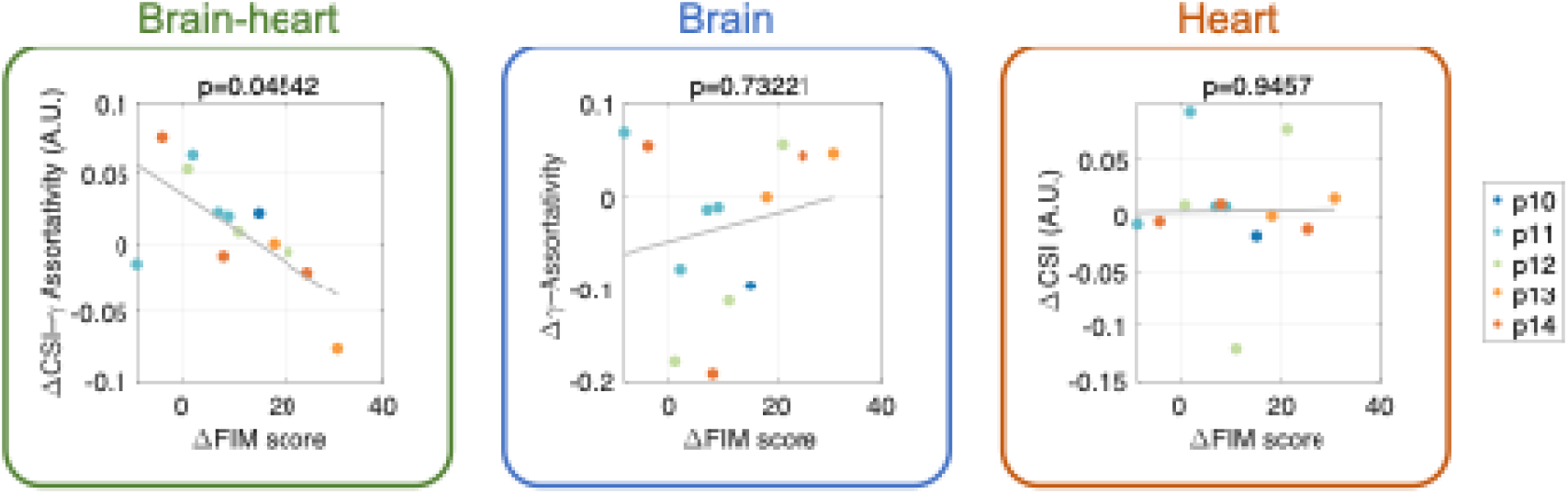
Changes in brain–heart interactions with respect to the changes in functional independence measures (FIM). The relationship was quantified from the coupling between gamma-band assortativity and cardiac sympathetic activity, in Dataset 2. Each of the 13 data points correspond to a change from one session to another for one of the 5 patients (p10 had 1 follow-up, p11 had 4 follow-ups, p12 had 3 follow-ups, p13 had 2 follow-ups and p14 had 3 follow-ups). A.U.: arbitrary units.

**Table 6.**
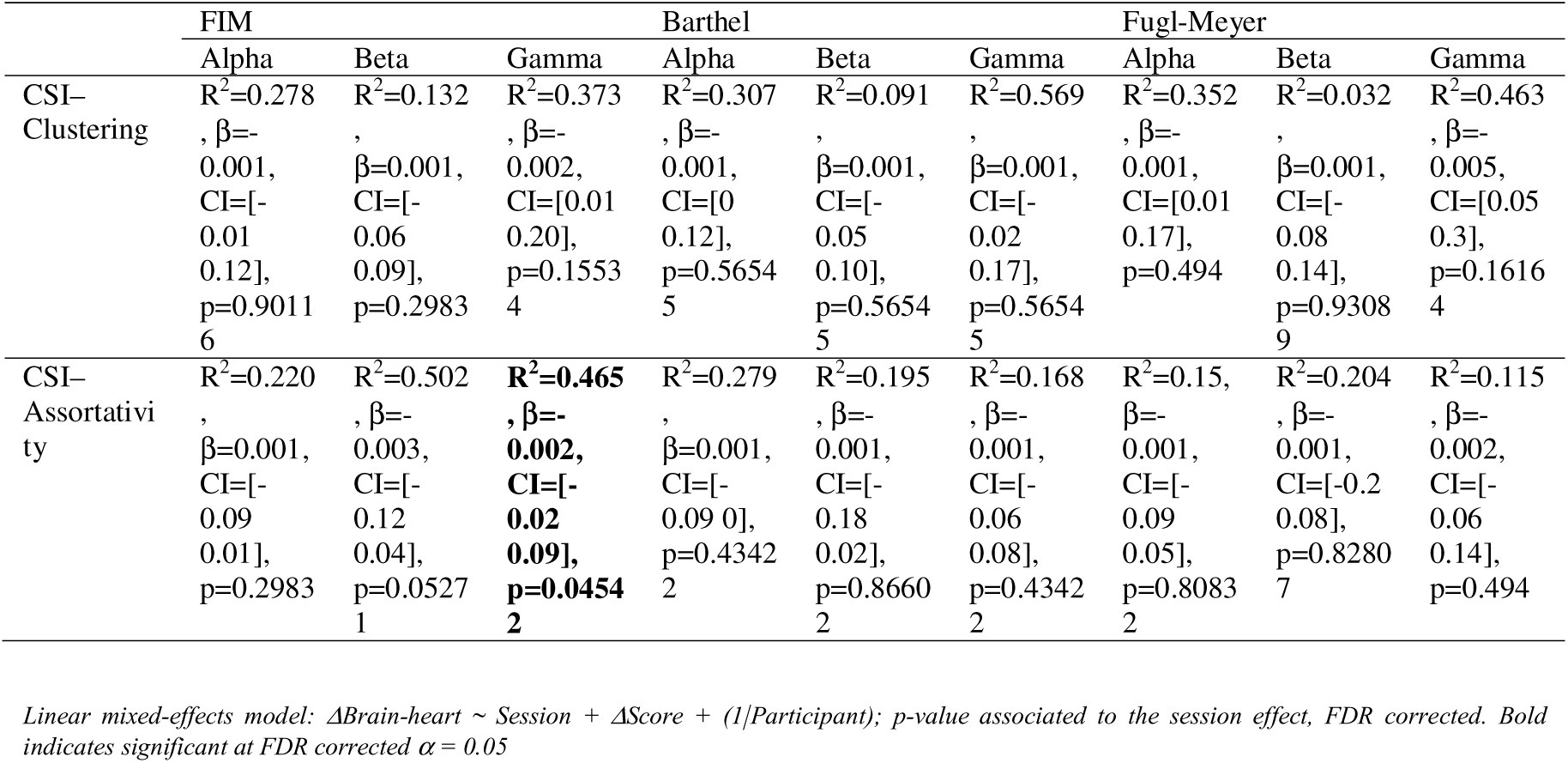
Linear mixed-effect model of the effect of clinical scores on brain-heart interactions during motor imagery.

**Table 7.**
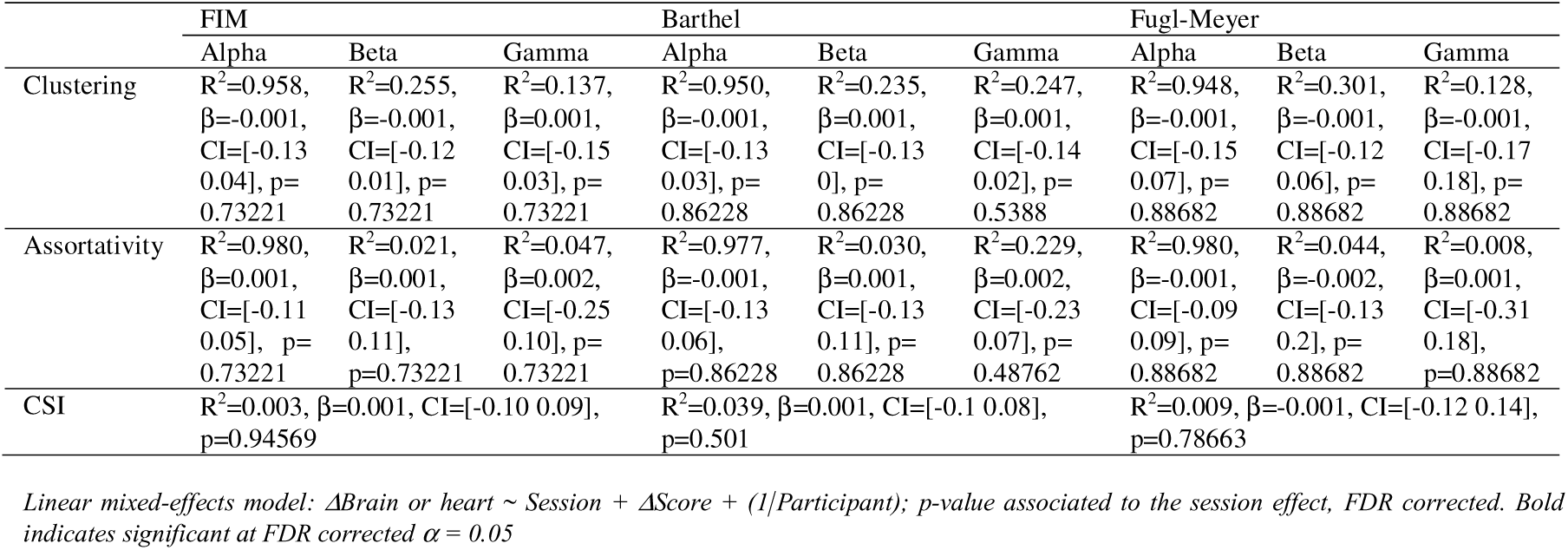
Linear mixed-effect model of the effect of clinical scores on brain or heart metrics during motor imagery.

## Discussion

In this study, we examined changes in brain–heart interactions during motor imagery interventions in stroke patients with either upper or lower limb motor impairments. Our findings revealed significant modulation of coupling patterns under two specific conditions: under electrical stimulation and in longitudinal interventions.

We observed a stronger association between cardiac sympathetic activity and brain network organization during motor imagery when electrical stimulation was applied to the affected limb. This effect spanned a wide range of EEG oscillation frequencies when electrical stimulation was synchronized with gait motor imagery, suggesting that synchronizing the task with stimulation enhances both task engagement and the related brain–heart dynamics. Notably, the association between cardiac sympathetic activity and gamma network organization remained robust even when stimulation was invariant. These results indicate that brain–heart interactions in the gamma band may serve as an index of engagement during motor imagery tasks, potentially providing a quantitative measure to capture interindividual variability—an aspect especially relevant in brain–computer interface applications and in patient populations with diverse etiologies. These results may also link to previous reports indicating that gamma activity serves as a predictor of motor imagery performance [54].

Regarding longitudinal changes in brain–heart interactions during motor imagery, patients in Dataset 1 (those with lower limb motor impairments and recruited during the chronic phase) showed a pronounced longitudinal effect across all brain–heart interaction metrics. Specifically, we found an increase in coupling between cardiac sympathetic activity and global brain dynamics, most prominently within the alpha–beta frequency range. In contrast, patients in Dataset 2, who were recruited during the acute phase and presented upper limb motor impairments, exhibited a more limited effect, restricted to the coupling between cardiac sympathetic activity and beta-band clustering. Beta-band activity is closely related to motor processes and their inhibition, and its network organization is known to be altered after stroke [1]. The observed effects may reflect a progressive refinement or stabilization of local processing within motor-related circuits [5,55–58], potentially supporting more efficient motor imagery over time. Gamma activity is often associated with local processing and integrative functions, and increased assortativity may indicate a reconfiguration of hub-to-hub interactions [40], enhancing the integration of information flow. The coupling of these dynamics with cardiac sympathetic activity suggests that autonomic processes may play an active role in shaping large-scale network reorganization during stroke recovery.

We then investigated the relationship between changes in clinical scores and brain–heart interactions during motor imagery. Overall, patients presented longitudinal improvements in clinical scores and longitudinal modulations in their brain–heart interactions. However, among the different clinical measures, only the Functional Independence Measure showed a significant association, specifically with changes in the coupling between cardiac sympathetic activity and gamma-band assortativity. This finding suggests that improvements in functional independence may be linked to more efficient integration of brain networks operating in the gamma frequency range, which are known to support sensorimotor processing and cognitive engagement. The modulation of cardiac sympathetic activity in parallel with gamma network reorganization could therefore reflect a more coordinated brain–heart response that accompanies functional recovery.

Sympathetic dynamics play a crucial role in multiple cerebral processes, including the maintenance of adequate cerebral perfusion through the modulation of heart rate, vascular tone, and cardiac output [59]. Although an intact autonomic state is recognized as essential for effective cerebral autoregulation [60], findings remain mixed regarding what aspects of cardiac autonomic control are truly reflected by heart rate variability analyses [28,61]. Following stroke, central autonomic pathways and cerebrovascular networks may be disrupted, leading to asymmetries in neurovascular control, particularly when lesions involve cortical or subcortical regions that mediate motor and autonomic integration [23].

By further uncovering the dynamic coupling between central and autonomic processes during motor imagery [19], our results open new avenues for targeted interventions and objective monitoring of their effects. In particular, these insights may inform the development of neuromodulation strategies aimed at simultaneously engaging central and autonomic pathways [32,62], an approach that has already shown promise in improving clinical outcomes after stroke.

This study has several limitations that should be acknowledged. First, the sample size was relatively small, comprising only 15 stroke patients with highly heterogeneous etiologies, lesion locations, and recovery trajectories. In addition, not all participants completed every experimental session, reflecting the inherent challenges of conducting longitudinal research with clinical populations prone to variability and dropout. Despite this heterogeneity, our findings revealed strong similarities between the two datasets, suggesting that the observed effects may extend beyond lesion location or the specific timing of the measurements. It is also important to note that our analyses were based on global brain network characterizations, which are lesion-location agnostic and enabled us to consider brain function at the whole-system level rather than focusing on localized damage. This approach may help generalize the observed brain–heart interactions across diverse stroke presentations.

Our results point to targeted interactions between specific frequency bands and network properties [6,40], suggesting that neuroplasticity following stroke involves finely tuned multisystem coordination. Overall, these findings support the view that brain–heart interactions are not merely a consequence or cardiac complications after stroke but may actively signal the reorganization of functional brain networks during the recovery of sensorimotor function. The progressive nature of the observed effects further suggests that these interactions could serve as sensitive biomarkers of neuroplasticity and recovery. Future work should investigate their relationship with behavioral improvements and explore their potential integration into brain–computer interface-driven rehabilitation protocols.

## Declarations

### Ethics approval and consent to participate

The study associated to the Dataset 1 was approved by the Ethics Committee of Tianjin Huanhu Hospital (IEC-B-013-V3.0) and followed the Declaration of Helsinki.

The study associated to the Dataset 2 was approved by the Ethics Committee CPP-IDF-VI of Paris and followed the Declaration of Helsinki.

### Consent for publication

Not applicable.

### Availability of data and materials

Dataset 1 is a publicly available resource [34]. Dataset 2 is available upon reasonable request to the corresponding author. To facilitate reproducibility, the source codes implementing the methods employed in this study are publicly available, including routines for cardiac sympathetic activity analysis [47], brain network metrics computation [43], and non-linear coupling estimation [53].

## Authors’ contributions

DCR: Conceptualization, Methodology, Software, Validation, Formal Analysis, Investigation, Resources, Writing Original Draft, Writing Review and Editing.

MCC: Data Curation, Investigation, Writing Review and Editing.

JGA: Data Curation, Writing Review and Editing.

CR: Data Curation, Investigation, Writing Review and Editing. FDVF: Investigation, Resources, Writing Review and Editing.

MC: Methodology, Investigation, Supervision, Writing Review and Editing. All authors read and approved the final manuscript.

